# Exome copy number variant detection, analysis and classification in a large cohort of families with undiagnosed rare genetic disease

**DOI:** 10.1101/2023.10.05.23296595

**Authors:** Gabrielle Lemire, Alba Sanchis-Juan, Kathryn Russell, Samantha Baxter, Katherine R. Chao, Moriel Singer-Berk, Emily Groopman, Isaac Wong, Eleina England, Julia Goodrich, Lynn Pais, Christina Austin-Tse, Stephanie DiTroia, Emily O’Heir, Vijay S. Ganesh, Monica H. Wojcik, Emily Evangelista, Hana Snow, Ikeoluwa Osei-Owusu, Jack Fu, Mugdha Singh, Yulia Mostovoy, Steve Huang, Kiran Garimella, Samantha L. Kirkham, Jennifer E. Neil, Diane D. Shao, Christopher A. Walsh, Emanuela Argili, Carolyn Le, Elliott H. Sherr, Joseph Gleeson, Shirlee Shril, Ronen Schneider, Friedhelm Hildebrandt, Vijay G. Sankaran, Jill A. Madden, Casie A. Genetti, Alan H. Beggs, Pankaj B. Agrawal, Kinga M. Bujakowska, Emily Place, Eric A. Pierce, Sandra Donkervoort, Carsten G. Bönnemann, Lyndon Gallacher, Zornitza Stark, Tiong Tan, Susan M. White, Ana Töpf, Volker Straub, Mark D. Fleming, Martin R. Pollak, Katrin Õunap, Sander Pajusalu, Kirsten A. Donald, Zandre Bruwer, Gianina Ravenscroft, Nigel G. Laing, Daniel G. MacArthur, Heidi L. Rehm, Michael E. Talkowski, Harrison Brand, Anne O’Donnell-Luria

## Abstract

Copy number variants (CNVs) are significant contributors to the pathogenicity of rare genetic diseases and with new innovative methods can now reliably be identified from exome sequencing. Challenges still remain in accurate classification of CNV pathogenicity. CNV calling using GATK-gCNV was performed on exomes from a cohort of 6,633 families (15,759 individuals) with heterogeneous phenotypes and variable prior genetic testing collected at the Broad Institute Center for Mendelian Genomics of the GREGoR consortium. Each family’s CNV data was analyzed using the *seqr* platform and candidate CNVs classified using the 2020 ACMG/ClinGen CNV interpretation standards. We developed additional evidence criteria to address situations not covered by the current standards. The addition of CNV calling to exome analysis identified causal CNVs for 173 families (2.6%). The estimated sizes of CNVs ranged from 293 bp to 80 Mb with estimates that 44% would not have been detected by standard chromosomal microarrays. The causal CNVs consisted of 141 deletions, 15 duplications, 4 suspected complex structural variants (SVs), 3 insertions and 10 complex SVs, the latter two groups being identified by orthogonal validation methods. We interpreted 153 CNVs as likely pathogenic/pathogenic and 20 CNVs as high interest variants of uncertain significance. Calling CNVs from existing exome data increases the diagnostic yield for individuals undiagnosed after standard testing approaches, providing a higher resolution alternative to arrays at a fraction of the cost of genome sequencing. Our improvements to the classification approach advances the systematic framework to assess the pathogenicity of CNVs.

## INTRODUCTION

Copy number variants (CNVs) are imbalances of genomic material compared with the reference genome resulting in the addition (duplications and insertions) or removal (deletions) of genomic segments. They vary in size but are defined as variants of more than 50 bp.^1,2^ CNVs are significant contributors to rare genetic disease.^3,4^ Chromosomal microarrays (CMA) have been the recommended first-line clinical test to investigate individuals with suspected rare genetic diseases, especially for multiple congenital anomalies and intellectual disability disorders,^5,6^ though practice is moving towards exome sequencing as a first-line test.^7^ Standard clinical CMAs can only detect CNVs larger than 50-100 kilobases, so this low resolution precludes most gene- and exon-level detection of CNVs. Due to technical challenges, CNVs have not traditionally been identified by standard exome sequencing which typically focuses on single nucleotides variants (SNVs) and indels.

Traditionally, exome-based CNV algorithms^8–10^ have relied on exome read depth to inform of the underlying copy number at a given locus. However, many factors influence exome read depth so detecting CNVs from exome data is difficult due to the non-uniform distribution of captured reads secondary to biases introduced by PCR amplification, exome capture, and mapping. These factors make it challenging to differentiate between a technical artifact and a bona fide CNV. The GATK-gCNV tool^11^ uses a probabilistic framework to infer rare CNVs from read depth data in the presence of these systematic biases. The performance of GATK-gCNV has been benchmarked with genome sequencing; it achieved 97% precision in detecting *de novo* CNVs captured by genome sequencing in 99 children from families with autism spectrum disorder and achieved more than 95% sensitivity for rare CNVs captured by genomes that span more than 4 exons, and more than 90% positive predictive value at all CNV sizes.^11^

We used the GATK-gCNV algorithm to call CNVs across the Broad Institute Center for Mendelian Genomics (Broad CMG) exome cohort, a research center within the Genomics Research to Elucidate the Genetics of Rare Diseases (GREGoR) consortium. The Broad CMG has performed exome sequencing on more than 6,000 families with a suspected genetic disease since 2016, representing a large cohort of individuals with heterogeneous phenotypes including neurodevelopmental disorders, neuromuscular diseases, retinal disorders, blood disorders, kidney diseases, multiple malformations syndromes, and other conditions. Most individuals have had prior gene panels, exome, and/or clinical CMA but the level of prior genetic testing is variable. Several molecular diagnostic laboratories and many research groups have incorporated CNV calling in their exome analysis, particularly in recent years. The reported additional diagnostic yield of CNV calling on exome data, most commonly used as a second-line test after CMA, on various cohorts of patients with suspected rare genetic diseases varies between 1 to 2%.^12–16^

The widespread implementation of CMA and exome/genome sequencing is expanding the types and numbers of CNVs identified in both clinical and research settings, and it can be challenging to determine the impact of these CNVs on human health. Several resources have been or are being developed to address this challenge. For instance, high quality reference population data such as gnomAD SV,^17^ a reference dataset of structural variants (SV) from short-read genome sequencing of 10,847 individuals from the general population, helps determine the frequency of a CNV in the population. Also, *in silico* prediction tools for CNVs are available including some that have been developed with the goal of helping to distinguish deleterious CNVs from non-deleterious CNVs. For example, the StrVCTVRE score is a predictive tool that incorporates gene importance, conservation, coding sequence, and exon structure of the disrupted region and can evaluate CNVs overlapping coding sequences.^18^ CADD-SV, another example, is a tool developed using machine-learning random forest models to differentiate deleterious from neutral SVs.^19^

Importantly, accurate classification of CNV pathogenicity requires a consistent and transparent approach to be used across the human genetics field. Riggs *et al.* developed the American College of Medical Genetics and Genomics (ACMG) and the Clinical Genome Resource (ClinGen) consensus standards to guide in the evaluation of germline CNVs and encourage consistency in CNV interpretation across laboratories, technologies and specialties.^20^ They proposed a quantitative evidence-based evaluation framework to classify copy number loss and copy number gain that follow an autosomal dominant inheritance. These standards did not intend to cover all curation scenarios and, for example, do not extend to guidance on how to score CNVs following an autosomal recessive or X-linked inheritance, CNVs with available functional evidence, or SVs beyond deletions and duplications. Here, we developed and applied additional evidence criteria to address these limitations and assess the pathogenicity of all CNVs that were thought to be causal in the Broad CMG exome cohort.

## METHODS

### Case selection

The Broad CMG was established in 2016 as part of an initiative funded by the National Human Genome Research Institute of the National Institutes of Health, with the goal of discovering the variants and genes underlying Mendelian disease to increase diagnosis rates for individuals with a suspected genetic condition.^21–23^ The Broad CMG is now part of the NHGRI Genomics Research to Elucidate the Genetics of Rare diseases (GREGoR) consortium, the focus of which includes evaluating different approaches to improve rare disease diagnosis, such as CNV calling on exome data. Families recruited and sequenced through the Broad CMG are enrolled in research studies with local institutional review board (IRB) approval, including for sharing de-identified samples for sequencing and analysis (MassGeneralBrigham 2013P001477). Phenotypic information for the affected individuals in each family was provided using HPO terms.^24^

From February 2016 to May 2021 (5 years, 3 months), 6,633 families underwent CNV calling on exome data through the Broad CMG (15,759 individuals). This cohort had heterogeneous phenotypes including neurodevelopmental, neuromuscular, multiple congenital anomalies, hematological, ocular or renal disorders. Most were enrolled due to an unrevealing prior genetic diagnostic evaluation as many had a CMA, gene panel sequencing for known causes of disease, or clinical exome prior to research exome through the CMG. The sequenced individuals were submitted from a large number of studies and had variable levels of pre-screening prior to enrollment (and this information was not systematically collected).

### Exome sequencing

Exome sequencing was performed by the Genomics Platform at the Broad Institute of MIT and Harvard. Libraries from DNA samples (>250 ng of DNA, at >2 ng/ul) were created with an Illumina Nextera exome capture (37 Mb target) and sequenced (150 bp paired reads) to cover >80% of targets at 20x and a mean target coverage of >80x from February 2016 through January 2019 and then using a Twist exome capture (38 Mb target) and sequenced (150 bp paired reads) to cover > 80% of targets at 20x and a mean target coverage of >60x thereafter. Sample identity quality assurance checks were performed on each sample. The exome data was de-multiplexed and each sample’s sequence data were aggregated into a single Picard CRAM file. The BWA aligner was used for mapping reads to the human genome build 38 (GRCh38). Single nucleotide variants and insertions/deletions (indels) were jointly called across all samples using Genome Analysis Toolkit (GATK) HaplotypeCaller package version 3.5.

Default filters were applied to SNV and indel calls using the GATK Variant Quality Score Recalibration (VQSR) approach. Annotation was performed using Variant Effect Predictor (VEP), during upload of the callset to seqr^25^ for collaborative analysis between the Broad CMG team and collaborating investigators.

### CNV detection on exome data

CNVs were detected from exome sequencing following GATK-gCNV best practices^11^, as follows: read coverage was first calculated for each exome using GATK CollectReadCounts. After coverage collection, all samples were subdivided into batches of a median of 410 samples (range:160-625) for gCNV model training and execution; these batches were determined based on a principal components analysis (PCA) of sequencing read counts. After batching, one gCNV model was trained per batch using GATK GermlineCNVCaller on a subset of training samples, and the trained model was then applied to call CNVs for each sample per batch. Finally, all raw CNVs were aggregated across all batches and post-processed using quality- and frequency-based filtering to produce the final CNV callset. Methods are further described in Babadi et al.^11^

### CNV analysis

Each family’s CNV data was manually analyzed in coordination with the SNV/indel data by members of the Broad CMG analysis team using our in-house developed analysis platform, *seqr,* an open-source, web-based tool for family-based monogenic disease analysis that enables variant filtration, annotation and prioritization in addition to data sharing of candidate disease genes (with variants and HPO terms) through the Matchmaker Exchange.^25^ CNVs were filtered based on their mode of inheritance, gCNV quality scores (QS) (QS>50; developer recommendations are QS>50 for duplications, >100 for deletions, and >400 for homozygous deletions, see Babadi *et al*^11^ for details), and their frequency in the Broad CMG callset. For autosomal dominant conditions, we filtered for CNVs with an allele frequency of <0.1% in the Broad callset, and used <1% for autosomal recessive conditions. When analyzing each family, factors used to help prioritize if a CNV was of clinical significance for a given individual included the CNV size, its structural consequences (predicted loss-of-function (LoF) variant, copy gain), its segregation pattern within the affected family, its frequency in the gnomAD-SV^17^ reference population database, the number and characteristics of genes involved in the CNV, and *in silico* prediction of pathogenicity tools. Of note, the following criteria needed to be met for a SV in gnomAD to be considered as the same allele:

– same SV type (duplication, deletion, etc)
– either has sufficient reciprocal overlap (50% reciprocal overlap for large SV >5Kb; 10% reciprocal overlap for SV <5Kb).

Genes included in a CNV were evaluated for gnomAD gene constraint scores, ClinGen dosage sensitivity scores and disease association in OMIM; exons included in an intragenic CNV were evaluated for exon expression (pext score in gnomAD^26^) and conservation. If no promising variants were found using our initial searches, we removed the QS filter to include low-quality variants. We reviewed the StrVCTVRE score^18^ of candidate CNVs but did not use it to filter data or rule out variants. The score ranges from 0-1, a score of 1 being more deleterious. In line with the developer suggestions, CNVs with a score >0.37 were considered as having a higher likelihood of being deleterious. To evaluate the quality of a given CNV, the patient’s copy number level was compared to any additional sequenced family members as well as a cluster of other samples with similar read depth that act as controls. The copy number plot of each compelling candidate was assessed to confirm an increase or decrease (corresponding to either a gain or a loss) between the proband and the background cluster, and a difference in the proband’s copy number within versus outside the reported coordinates of the CNV (Figure 1).

**Figure 1.**
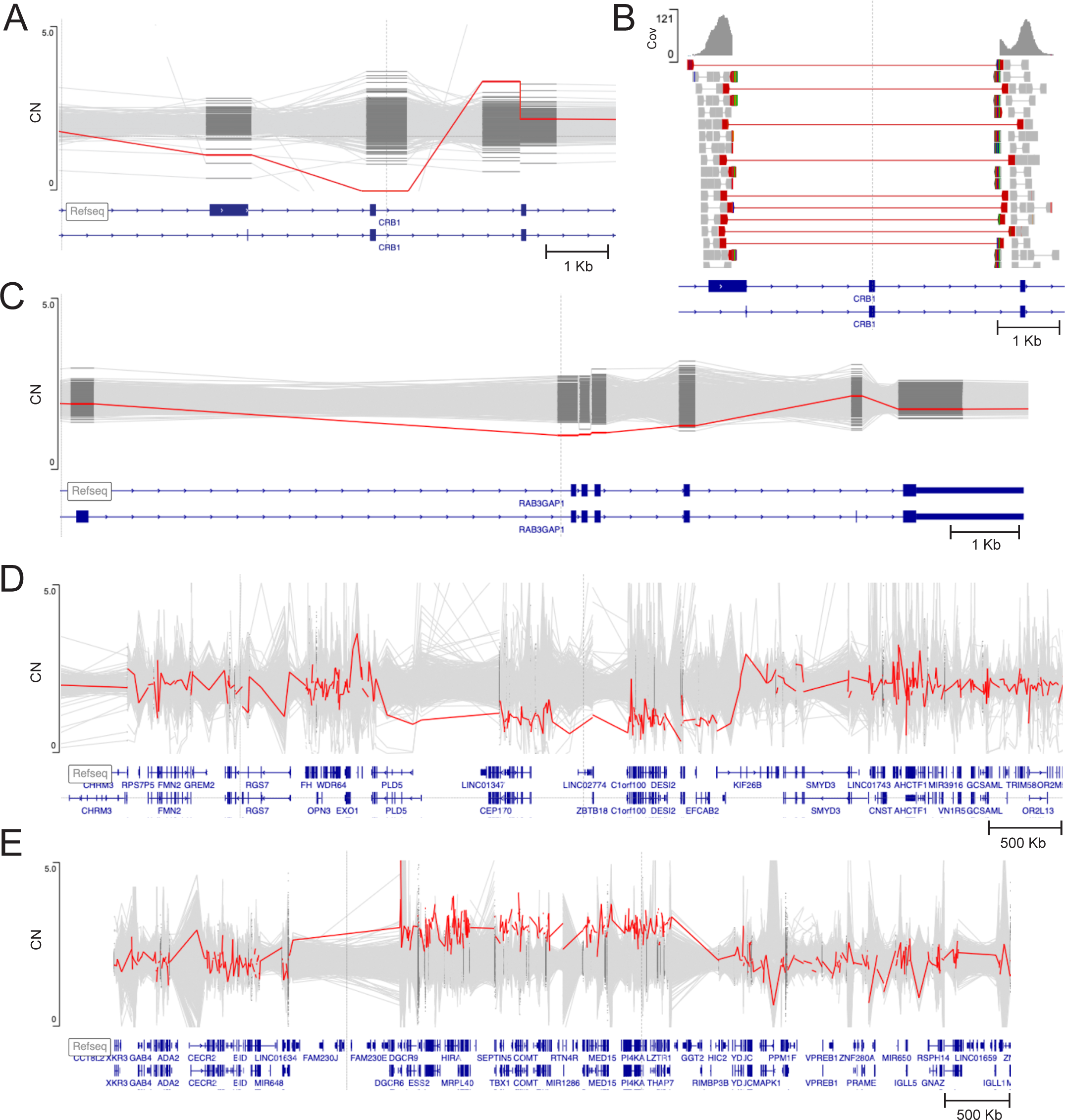
Exome copy number plot and reads visualization for examples of causal CNVs in the Broad CMG cohort. (A) Individual affected with retinitis pigmentosa with a homozygous single exon deletion in *CRB1* (chr1:197438450-197439442×0, Quality score (QS) = 120) identified on exome. To evaluate the quality of the CNV, the patient’s copy number (CN) level (in red) was compared to a cluster of other samples with similar read depth that act as controls. The proband’s CN is decreased compared to the background cluster, compatible with a homozygous deletion. Y axis: CN. (B) As breakpoints fell within the exome data, manual inspection of read data from the individual from (A) using the Integrated Genomics Viewer (IGV) showed discordant read pairs, split reads and complete absence of coverage, compatible with a homozygous exon 10 deletion also including part of upstream exon 9 in *CRB1* (chr1:197435257-197441674×0 (NM_201253.3)). Cov= coverage. (C) Individual with multiple congenital anomalies and a heterozygous deletion of 4 exons in *RAB3GAP1* (Warburg micro syndrome) (red, chr2:135162318-135164794×1, QS =92) in trans with a frameshift variant in *RAB3GAP1* (not shown, NM_012233.3: c.2393_2394del, p.Leu798ArgfsTer7), both identified by exome. The presence of the deletion was validated by droplet digital PCR. Y axis: CN. (D) Individual with a neurodevelopmental disorder with a *de novo* 2.6 Mb heterozygous 1q43q44 deletion (red, chr1:242523991-245156781×1, QS =3077) identified on exome. The presence of this deletion was validated by quantitative PCR. Y axis: CN. (E) Individual with a neurodevelopmental disorder with a *de novo* 2.1Mb 22q11.2 duplication (red, chr22:18985739-21081116×3, QS =3077) identified on exome. The presence of this duplication was validated by chromosomal microarray. Y axis: CN. All coordinates on GRCh38.

We also visually inspected the read data of candidate CNVs using the Integrated Genomics Viewer (IGV) to evaluate for sequencing artifacts (Figure 1).

A CNV is defined as high-confidence by GATK-gCNV (see Babadi *et al.*^11^ for details) if:

● The CNV is present in a high-quality sample (with ≤ 200 autosomal raw CNV calls, of which at least 35 have QS >20)
● The sample frequency of the call is ≤ 0.01 within the Broad callset
● The number of overlapped exons is ≥ 3
● The QS score is equal or greater than the QS threshold (QS>50 for duplications, >100 for deletions, and >400 for homozygous deletions)

### CNV validation

CNV validations were performed by the investigator that contributed the sample by a variety of methods (FISH, karyotype, CMA, MLPA, Sanger sequencing, quantitative PCR, droplet digital (dd)PCR^27^, genome sequencing) across different clinical or research laboratories, while some were validated by short read or long read genome sequencing performed at the Broad Genomics Platform (Table S1). Not all CNV identified by the gCNV pipeline were validated by another method, largely when samples were from historic cohorts where there was not a path to return results and often insufficient remaining DNA.

### Evaluation of CMA coverage for each causal CNV

To evaluate how many causal CNVs could have been detected by a standard clinical CMA, CNV detection sensitivity by CMA was assessed by evaluating the number of probes from the Agilent GenetiSure Cyto CGH+SNP arrays (downloaded from https://genome.ucsc.edu/ on May 23, 2023) included within the genomic coordinates of a given CNV. A minimum number of five probes was required to consider that the CNV would confidently be called by CMA^28^.

### Assessment of the pathogenicity of CNVs

We considered a case solved if the CNV was classified as pathogenic or likely pathogenic and conclusively explained the phenotype or if a variant was found involving a novel disease gene with moderate/strong supporting evidence by the ClinGen gene-disease validity criteria.^29^ Supporting genetic and/or experimental evidence were required to consider a CNV in a novel gene as the diagnosis in a given family, most often by additional families identified through Matchmaker Exchange. We also considered a case solved for cases where the analysis team and referring provider, when relevant, considered the variant causative, even if a CNV was technically a variant of uncertain significance (VUS) by ACMG/ClinGen CNV criteria.

Each CNV was evaluated and classified by two curators (GL and KR). In order to systematically assess the pathogenicity of the SVs in this study, the ACMG/ClinGen standards for interpretation and reporting of constitutional copy-number variants were applied.^20^ For candidate novel disease genes, the interpretation of gene-disease relationship was guided by the ClinGen framework.^29^ We developed an approach, including new curation criteria, to optimally capture evidence for pathogenicity for the range of variants discussed in this article.

### Determination of the number of protein-coding genes included in a CNV

In order to score points from section 3 from the Riggs standards (“evaluation of gene number”), we used OMIM gene number count (https://genescout.omim.org/), and have compared it to the gene number count provided by the DECIPHER browser (https://www.deciphergenomics.org/) and the ClinGen browser (https://search.clinicalgenome.org/kb/gene-dosage?page=1&size=25&search=).

### Variants following autosomal recessive inheritance

The current ACMG/ClinGen CNV standards do not yet provide guidance on how to score CNVs in genes for conditions that follow an autosomal recessive inheritance. To classify these variants within this project, we developed an approach, advancing the current framework.

● We applied category 2E and the PVS1 LoF flowchart^30^ for any intragenic CNV, or if a CNV had a complete or partial overlap with a gene with an established gene-disease relationship that follows an autosomal recessive inheritance.
● When the candidate CNV involved a gene with no established gene-disease relationship, we did not score points from category 2, but rather used category 4 to build up evidence for an established gene-disease relationship by finding additional cases with overlapping variants from the literature.
● Points were awarded to the Broad CMG cases and published cases from the literature

using a similar system to that which is used when curating SNVs (the PM3 criteria) [ClinGen Sequence Variant Interpretation Recommendation for in trans Criterion (PM3) - Version 1.0 Working Group Page: https://clinicalgenome.org/working-groups/sequence-variant-interpretation/, Approved: May 2, 2019]. The point-based system suggested in the PM3 criteria was translated into points of similar strength level in the Riggs quantitative framework^20^ (Table 1).

- ● We added 0.15 points when at least one individual with a unique phenotype (phenotype is highly specific to disease, low genetic heterogeneity) has been reported by our study or in the literature (equivalent of PP4 criteria in Richards et al^31^)
- In some cases, we awarded 0.30 points when evidence was particularly strong.

**Table 1.** Adapted PM3 table to score CNVs in genes for conditions that follow an autosomal recessive inheritance.

| Variant classification/zygosity | Points per Proband |  |
| --- | --- | --- |
|  | Confirmed in *trans* | Phase unknown |
| Second variant is pathogenic (P) or likely pathogenic (LP) | 0.30 | 0.15 (P)<br>0.08 (LP) |
| Homozygous occurrence of this variant<br>(*max 0.30 point*) | 0.15 | N/A |
| Second variant is a variant of uncertain significance<br>(*max 0.16 point*) | 0.08 | 0.0 |

This only applied for genetic diseases with a specific, unique phenotype, high clinical sensitivity testing (e.g. biochemical assays, enzyme deficiency assays, functional cytogenetic tests (e.g. chromosomal breakage study)), and consistent family history. These additional points were only used one time per variant.

### Variants following X-linked inheritance

We developed the following flowchart to score points for CNVs with X-linked inheritance (Figure 2).

**Figure 2.**
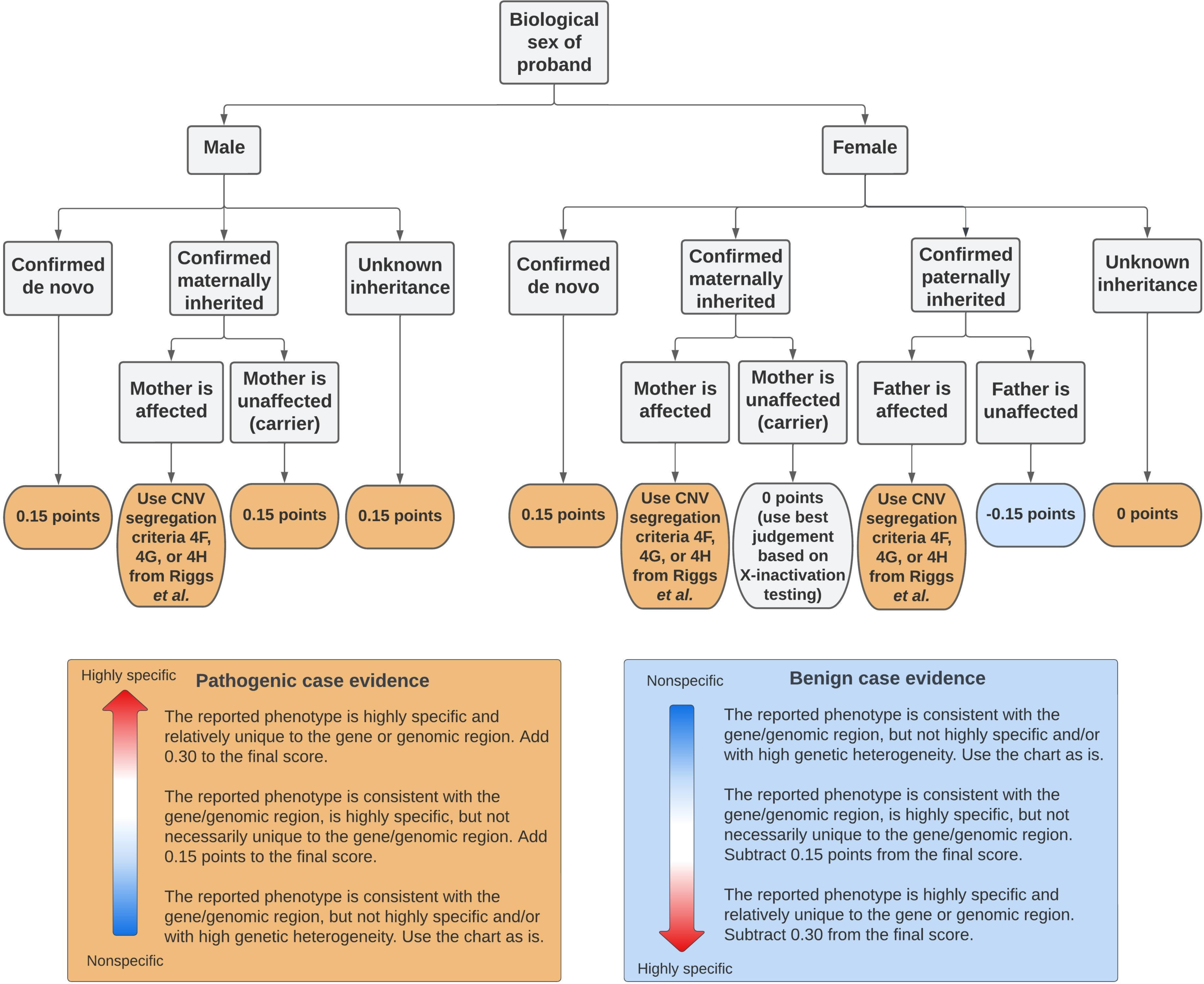
Flowchart illustrating how points were scored for CNVs that followed a X-linked inheritance. We incorporated sex of proband, parental genotype and parental affected status to score both the proband in which the X-linked variant was identified and, if applicable, any individual in the published literature or public databases that had variants of similar genomic content to the variant of interest. The points for each case could be increased or decreased based on phenotype specificity, up to 0.45 points.

### Complex SVs

We defined a complex SV as a complex rearrangement typically composed of three or more breakpoint junctions that cannot be characterized as a single canonical SV type^32^. Some complex SVs were suspected on exome CNV analysis and/or identified after further validation. As suggested by Riggs *et al.*^20^, when classifying complex rearrangements (for example a paired duplication inversion), we evaluated each CNV separately. The overall classification for the event was defaulted to the most deleterious classification (for example, if the deletion portion were classified as “pathogenic” and the duplication portion was classified as “uncertain significance,” the entire SV was classified as “pathogenic”).

### Inversions and Insertions

For variants initially called as deletion or duplication by GATK-gCNV in this cohort, some were identified as including inversions or insertions by validation methods. The Riggs *et al.*^20^ standards do not provide guidance on how to score inversions or insertions. Therefore, we took guidance from Collins *et al.*^33^ which states that inversions can be evaluated as a LoF event if exactly one breakpoint falls within a gene, or both breakpoints fall within the same gene and span at least one exon. Collins *et al.* also recommend evaluating a large insertion within an exon as a LoF event. We applied the LoF PVS1 criteria^30^ as appropriate for such cases.

### Variants with available functional evidence

We added an additional 0.15 points for any variant with at least supporting functional evidence of pathogenicity, either from the investigation of our cases or from the literature. Examples included: expression assays (Western blot for protein expression, PCR for RNA expression), RNA sequencing, cellular assays (impaired localization and/or function) or protein interaction studies. If the evidence was stronger, the points were upgraded to “moderate” (0.30 points) or strong (0.45 points). For example, RNA sequencing results showing a clear and significantly decreased expression of a gene or an animal model with the exact variant recapitulating the disease phenotype was given 0.45 point (strong evidence).

## RESULTS

CNV calling using the GATK-gCNV algorithm was performed on exomes from the Broad CMG cohort of 6,633 families with heterogeneous rare disease phenotypes and variable prior genetic testing that typically included a gene panel, exome, and/or CMA. A total number of 9,930 high-confidence (as defined in Babadi *et al.*^11^) unique variants (4,387 deletions and 5,543 duplications) were identified across 15,759 individuals from these 6,633 families (Figure 3A and Figure S1), 10,472 of the 15,759 samples had at least one rare (<1% frequency in the Broad data callset) high-confidence CNV, and the median number identified was two (sd+-1.55) per individual (Figure S1). The entire CNV callset for these individuals, with a total of 2,131,645 copy number calls (292,833 unique variants), was loaded into the seqr platform for analysis.

**Figure 3.**
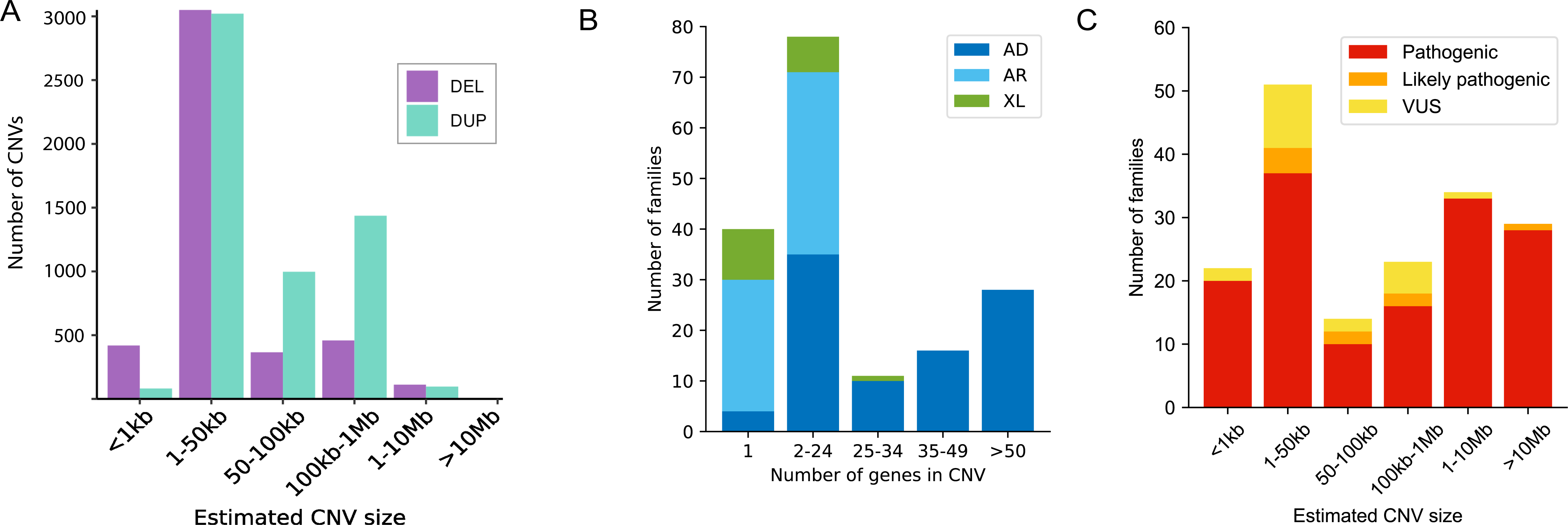
Characteristics of CNVs across the entire callset and the subset of causal CNVs. (A) Number of high-confidence CNVs by estimated size that were identified in the Broad CMG exome callset of 6,633 families sequenced between 2016 and 2021. Large CNVs tend to be fragmented into multiple small GATK-gCNV calls, accounting for why there are no CNVs in the >10 Mb category of the graph. These CNVs were interpreted as being part of the same underlying event when looking at the copy number plot and/or validation methods and are presented that way in Figure 3B and 3C. DEL: deletion; DUP: duplication. (B) Mode of inheritance and number of genes involved in each CNV in 173 families in which the CNV was interpreted as causal. The number of genes included in each interval was chosen based on cutoffs suggested for CNV scoring in section 3 of the Riggs *et al.* ACMG/ClinGen standards.^20^ (C) CNV classification by estimated size in 173 families in which the CNV was interpreted as causal by the multidisciplinary team. The causal CNVs consisted of 141 deletions, 15 duplications, 3 insertions (miscalled as deletion by GATK-gCNV), and 14 complex structural variants (SV). We interpreted 153 CNVs as likely pathogenic/pathogenic and 20 CNVs as VUS.

Many of these low-quality calls were likely artifacts, but by incorporating phenotype and allelic variation (SNVs, indels, CNVs) in the analysis of each family, some low-quality CNV calls were prioritized and ultimately interpreted as causal. Through the entire callset analysis, we have identified a causal variant in 173 previously undiagnosed families. CNV calling on existing exome data in this cohort thus resulted in an additional solve rate of 2.6% (173/6,633). The causal CNVs consisted of 144 deletion, 15 duplication, and 14 suspected complex (multiple CNVs on a chromosome) GATK-gCNV calls, which are currently resolved as 141 deletions, 15 duplications, 3 insertions, 10 complex SVs and 4 suspected complex SVs. Of the 10 validated complex SVs, three were initially deletion or duplication calls where a complex SV was identified on validation.

These CNVs mostly involved established genes/loci, but five families that are considered solved had a CNV involving a novel disease gene candidate. Supporting genetic and/or experimental evidence was required to consider a CNV in a novel gene as the explanation for a given family, most often by additional families identified through Matchmaker Exchange^34^ or the literature. The disorder followed an autosomal dominant inheritance in 93 families, an autosomal recessive inheritance in 62 families and X-linked inheritance in 18 families (Figure 3B). The CNV was confirmed *de novo* in 70/93 (75%) of the families with an autosomal dominant disorder, inherited from a parent in 3/93 families (one inherited from an affected parent, one involving an imprinted locus, and one inherited from an unaffected parent for a condition known to harbor incomplete penetrance/variable expressivity) and the inheritance was unknown in 20/93 families. The CNV was confirmed *de novo* in 7/18 (39%) of the families with an X-linked disorder. Detailed information on the CNV of each family is provided in Table S1. The predominant phenotype present in the 173 families was neurodevelopmental disorders (54%) followed by neuromuscular disorders (15%), but the cohort with causal CNVs also included individuals with multiple congenital anomalies, hematological, ocular, and renal phenotypes.

The degree of prescreening before research exome differed between individuals from different sub-cohorts and was therefore non-uniform across different phenotypes. The estimated sizes of causal CNVs by exome ranged from 293 bp to 80 Mb (Figure 3C). Twenty-two CNVs involved one exon and 14 CNVs involved two exons, which is below the benchmarked resolution of GATK-gCNV indicating it may be able to detect even smaller CNVs when allowing for a higher false positive rate. Large CNVs were also identified as some individuals did not have CMA prior to research enrollment. Large CNVs tend to be fragmented into multiple small GATK-gCNV calls. We interpreted fragmented CNVs as being part of a larger CNV event in 35 families (35/173 (20%)) in this cohort after looking at the copy number plot and/or validation methods.

We sought to evaluate how many of the causal CNVs could have been detected by one of the standard clinical CMAs, which is distinct from a high-density clinical array which often has one or more probes per exon. Standard CMAs usually detect CNVs larger than 50-100 Kb but the resolution varies across the genome and across different array designs as the probes are not evenly spaced but are clustered around regions of clinical interest. CNV detection sensitivity by a representative standard CMA was assessed based on the minimum number of probes considered “sufficient” for CNV calling per target, which is defined as ≥5 probes for the Agilent GenetiSure Cyto array.^28^ Based on this, we estimate that 44% (76/173) of these CNVs are unlikely to have been detected by standard CMA.

More than half of the CNVs (105/173 (61%)) were validated by various orthogonal methods, such as CMA, PCR, FISH, karyotype, MLPA, Sanger across the CNV or breakpoints, or short or long read genome sequencing. Of note, some of these methods did not provide breakpoints but rather only confirmed the copy number change. Of the 105 validated CNVs, 30 (29%) showed differences when comparing the initial results with the orthogonal validation results: 19 showed differences in gene/exon content and 11 showed differences in SV type. Importantly, the difference in gene or exon content identified in 19 families did not result in a change in the clinical interpretation of the CNV. Of note, only one of these 19 CNVs was curated as a VUS and the difference in the number of exons included in the CNV did not change the scoring and classification of this CNV. The 11 cases with different SV type consisted of eight complex SVs which were either incompletely characterized or not suspected by GATK-gCNV on the exome, and a recurrent Alu insertion in the *MAK* gene (OMIM #614181)^35^ identified in three individuals with retinitis pigmentosa. This insertion was miscalled as a deletion by the GATK-gCNV pipeline, but manual inspection of the exome reads showed discordant read pairs compatible with an Alu insertion. Sanger sequencing resolved the nature of this event.

Overall, there were 10 confirmed complex SVs in this cohort. We defined a complex SV as a complex rearrangement typically composed of three or more breakpoint junctions that cannot be characterized as a single canonical SV type.^32^ A complex SV was suspected on the GATK-gCNV calls in 11 families (del/dup, paired dup, etc); seven of these were confirmed by genome, qPCR or CMA (Table S1) and four remained unvalidated. Two deletions and one duplication identified by GATK-gCNV in three different families were revealed to be complex SVs (paired deletion inversions and a paired inversion duplication) when validated by genome sequencing or long-range PCR.

Twenty-four unrelated families with causal CNVs had a recurrent CNV that was identified in more than one other unrelated family in this cohort. The recurrent 22q11.2 microdeletion syndrome (OMIM #188400) was identified in nine individuals with neurodevelopmental disorders in this cohort. Two individuals with a neurodevelopmental disorder were diagnosed with 22q13.3 deletion syndrome (Phelan-McDermid syndrome (OMIM #606232)). The 17q12 deletion syndrome (OMIM #614527) was identified in two individuals with renal cystic disease. There were multiple recurrent CNVs identified in the subgroup of individuals with retinal disorders in this cohort. Indeed, four individuals of European ancestry affected with cone rod dystrophy had a heterozygous 1-exon-deletion in *CLN3* (OMIM #204200) in trans with a pathogenic variant.^36^

A founder variant in the Ashkenazi Jewish population, an Alu insertion in *MAK* (OMIM #614181)^35,37^, was found in three affected individuals of this ancestry. Two individuals of different ancestries affected with retinitis pigmentosa were homozygous for the same 2-exon-deletion in *EYS* (OMIM #602772), a deletion previously reported in the literature.^36,38^ Two individuals of European ancestry affected with retinitis pigmentosa had a heterozygous 4-exon-deletion in *EYS* (OMIM #602772), a deletion reported in multiple affected individuals in the literature ^36,39–41^ in trans with a pathogenic or likely pathogenic variant. Detailed information on the CNV of each of these families is provided in Table S1.

The StrVCTVRE *in silico* score was evaluated across the cohort. This score was viewable on each CNV within seqr during the initial analysis but was not used for filtering and not strongly relied on in analysis (consistent with how other *in silico* scores are viewed in our analysis pipeline). Sharo *et al.* reported that a 90% sensitivity is reached at a StrVCTVRE score of 0.37 (score ranges from 0-1, a score of 1 being more deleterious), which suggests that when used on a collection of SVs called from a clinical cohort, this threshold may identify 90% of pathogenic SVs while reducing the candidate SV list by 54%.^18^ In this cohort, 161/168 unique causal CNVs had a StrVCTVRE score greater than 0.37 (true positive rate of 0.96%), while this was the case for 6162/10788 non-causal CNVs (false positive rate 0.57) (Table S2). The median score of the 161 unique causal CNVs was 0.77, and 0.42 for non-causal CNVs that had a StrVCTVRE score calculated. One minor limitation of this analysis is that many large CNVs are fragmented, which may result in lower StrVCTVRE scores for constituent parts than would be assigned for the larger CNV event. While we manually reassembled and recalculated StrVCTVRE scores for causal CNVs reported here (as it is appropriate to apply these scores to the entire CNV), non-causal CNVs were not reassembled. We note that all CNVs greater than 3Mb size automatically had a score of 1 demonstrating a correlation between the CNV size and the StrVCTVRE score (Figure S2).

Using the 2020 ACMG/ClinGen CNV interpretation standards^20^ and additional evidence criteria that we developed (detailed in the Methods section), we interpreted 153 CNVs as likely pathogenic/pathogenic and 20 CNVs as VUS of high interest, including the 5 in novel disease-gene candidates (Figure 3C). When evaluating the pathogenicity of each CNV, we determined the number of protein-coding genes included in each CNV and compared that number to three different reference databases: OMIM (https://genescout.omim.org/), DECIPHER browser (https://www.deciphergenomics.org/browser) and ClinGen browser (https://search.clinicalgenome.org/kb/gene-dosage?page=1&size=25&search<u>=</u>) (Table S1). The vast majority of CNVs (148/173 (86%)) showed differences in gene number between these three commonly used databases). Using the 2020 ACMG/ClinGen CNV interpretation standards^20^, different points are scored based on the number of genes included in a CNV (section 3 of the standards). For example, 0 points are given for a deletion with 0-24 genes, 0.45 points for a deletion of 25-34 genes, and 0.9 points for a deletion of more than 35 genes.

For copy gain, 0 points are given for 0-34 genes, 0.45 points are given for 35-49 genes and 0.9 points for more than 50 genes. We used the number of genes provided by the OMIM database to perform the curation. Using the OMIM database versus DECIPHER resulted in a different final score for 24/148 (16%) CNVs, but this would only alter the final classification for one CNV, as points were awarded from other sections. That altered case was a 857kb *de novo* 22q13 duplication which would be classified as a VUS if we use the gene number provided by OMIM (28 protein-coding genes) but would be classified as pathogenic if we had used DECIPHER browser (35 protein-coding genes). Detailed information on the CNV curation of each family is provided in Table S1.

## DISCUSSION

We present the analysis and curation results from CNV calling on exome data across a large and phenotypically heterogeneous cohort. The additional 2.6% solve rate of exome CNV calling identified in this cohort is comparable to previously reported diagnostic yield in other cohorts.^12–16^ In this cohort, most causal CNVs were deletions. Duplications were more common in the callset, but are less likely to disrupt gene function and also typically require more functional investigation to confirm a deleterious effect. Our callset contains many candidate duplications (and deletions) that could potentially elucidate additional affected families, but their pathogenicity remains uncertain and has not been further investigated.

Similar to using the probes on a microarray to estimate CNV size, the size of a CNV from exome analysis is an estimate based on which exons have an abnormal copy number, but the breakpoints typically occur somewhere within the introns. In addition, some exons have more heterogeneous coverage and the deletion or duplication may involve more or fewer exons than predicted. This can also result in a large CNV being called as multiple smaller events, but when the data is reviewed, it can often be assembled into a larger event.

In this study, we did not attempt to validate and map all the CNV breakpoints, and we did not assess the validation rate of GATK-gCNV as this has been done previously.^11^ A small number of CNVs were nonetheless confirmed by genome sequencing by the Broad CMG as part of initial efforts to validate gCNV performance. Of the 23 deletions and two duplications identified by GATK-gCNV and validated by Broad CMG genomes, these were resolved as 22 deletions, one duplication and two complex SVs. We recommended that any candidate CNV variants be confirmed with an orthogonal method and generally validations are performed by the collaborating researcher who recruited the individual for sequencing. The sensitivity of GATK-gCNV decays greatly for CNVs smaller than three exons (e.g. only ∼50% for CNVs involving 1 exon), but the precision is relatively stable^11^; interestingly, 36 CNVs (36/173 (21%)) in this cohort involved fewer than 3 exons, highlighting the benefit of reviewing the full dataset with the context of the patient’s phenotype and for some cases, a pathogenic variant in trans, can highlight small or poor quality CNV calls that warrant further attention. More than half of CNVs were validated by various methods and validation is either underway or may not be possible for the remainder of the identified CNVs. Importantly, the difference in size and in gene/exon content for validated CNVs did not lead to a change in the interpretation of any of the CNVs initially identified as causal, but it is possible that some interesting CNVs in this cohort were overlooked for that reason.

GATK-gCNV can only call deletions or duplications on exome data, so seven suspected complex SVs and three initially unsuspected complex SVs in this cohort were identified by orthogonal validation methods. We likely underdetected complex SVs as 39% of the CNVs in this cohort were not validated and some validation methods would miss a more complex event, such as droplet digital or quantitative PCR which only confirm the abnormal copy number without mapping the breakpoints.

There are only a few *in silico* prediction tools available for CNV interpretation. Our group used StrVCTVRE scores and we observed it was a useful tool to consider when prioritizing CNVs in this cohort. Generally, we use *in silico* predictions as accessory annotations for review when considering a variant rather than using it to filter out variants, even more so because large CNVs may be represented by multiple smaller fragmented calls. More data on analysis of cohorts of patients with rare diseases is needed to determine its utility overall and comparison to other available SV predictors. Of note, StrVCTVRE only provides a prediction score for CNVs overlapping a coding region, which was not a factor for this cohort given it was exome-based, but this is a limitation of the score when considering genome sequencing and noncoding SVs.

High-quality reference population data is essential for effective CNV analysis. The gnomAD SV database stands as a pivotal resource in human genetics but is currently limited to sequencing data from short-read genomes. We used the database to evaluate if a given CNV was present in the general population, which we found was useful for variant analysis and prioritization. There are a myriad of technical differences between genome and exome sequencing and, while studies have shown high overlap between CNV calling between the two techniques, the planned addition of CNV calling on gnomAD exomes is anticipated to improve clinical CNV interpretation since they will be more analogous from a technical standpoint. As the gnomAD SV dataset expands in terms of size (incorporating both exome and genome data) and ancestral diversity, its utility as an invaluable tool for both rare disease diagnosis and broader genetic studies will only increase.

Standards for CNV classification are an important yet challenging area requiring ongoing development. We proposed new evidence criteria to enable the assessment of the pathogenicity of all CNVs that were thought to be causal in our cohort. We identified four areas that needed additions or refinements. First, we suggested that functional data, including expression assays (Western blot, PCR, RNA sequencing) and cellular assays (localization/function), be incorporated as evidence at the supporting level of 0.15 points, and could be increased in weight as appropriate. For example, abnormalities observed in RNA sequencing data or an animal model with the same variant recapitulating the phenotype could be scored 0.3 or 0.45 points, respectively. Given the increasing availability of RNA sequencing, we suggest that incorporating scoring for functional evidence is essential for CNV classification. Second, to score CNVs involving genes associated with disorders with autosomal recessive inheritance, we proposed an approach inspired by the ACMG/AMP criteria PM3 used for SNVs by incorporating phase and classification of the second variant (Table 1). The point-based system suggested in the PM3 criteria was translated into points of similar strength level in the Riggs quantitative framework. We also used the PVS1 flowchart^30^ (or criteria 2E in Riggs *et al.*^20^) for intragenic CNVs or CNVs including at least one gene that had an established gene-disease relationship following an autosomal recessive inheritance. Additional points were added based on phenotype specificity and familial segregation. Third, to classify CNVs that follow an X-linked inheritance pattern, we developed a scoring system based on biological sex of the proband, parental genotype, and affected status of the transmitting parent (Figure 2). Points were upgraded by one or two strength levels based on phenotype specificity. We also used the PVS1 flowchart^30^ for intragenic CNVs or CNVs including at least one gene that had an established gene-disease relationship following an X-linked inheritance. Finally, to evaluate SVs other than deletion and duplication, we took guidance from Collins *et al.*^33^, which states that LoF can be expected if there is an insertion within an exon, if an inversion breakpoint falls within a gene, or if both inversion breakpoints fall within the same gene and span at least one exon. We thus applied the PVS1 LoF flowchart here. Our approach refined multiple aspects of CNV classification and advanced the systematic framework to assess the pathogenicity of CNVs.

An important step in CNV classification involves determining the number of protein-coding genes it contains. We observed some significant differences in gene number in CNVs evaluated in this cohort depending on which database was queried, the OMIM database being the most conservative. OMIM’s gene count results from manual curation of published references, while DECIPHER extracts this information directly from the Ensembl GRCh38 genome. OMIM might thus underestimate the real number of genes present in a CNV and DECIPHER might overestimate it. Even though different points were scored for several CNVs, the choice of which database to use did not affect the final classification except for one duplication in this cohort. For that duplication, the genes that were missing in OMIM but included in DECIPHER consisted of seven protein-coding genes. Our group opted for a conservative approach and used the OMIM database but this question needs to be further studied as this can lead to confusion during the curation process. In addition, a sliding scale to score progressive points based on the increasing number of genes in a given CNV could be used instead of fixed cutoffs, and features such as loss of function constraint, haploinsufficiency, and triplosensitivity scores could be incorporated.

## CONCLUSION

CNV calling and analysis from existing exome data increases the solve rate by 2.6% in this diverse and presumed monogenic cohort. This is a higher resolution alternative to arrays at a fraction of the cost of genome sequencing and can be applied retrospectively to existing exome datasets. We estimate that 44% of the 173 causal CNVs may not have been detected by standard clinical CMAs. In classifying these variants, we advanced the current standards to take into account additional types of evidence contributing to the systematic framework to assess the pathogenicity of CNVs.

## Data and code availability

The CNVs that were interpreted as causal in this cohort were submitted to ClinVar (https://www.ncbi.nlm.nih.gov/clinvar/) (submitter ID 506627, Broad Rare Disease Group). The ClinVar accession numbers of each CNV are listed in Table S1.

## Data Availability

All data produced in the present work are contained in the manuscript. The CNVs that were interpreted as causal in this cohort were submitted to ClinVar (https://www.ncbi.nlm.nih.gov/clinvar/) (submitter ID 506627, Broad Rare Disease Group). The ClinVar accession numbers of each CNV are listed in Table S1.

## Acknowledgements

We thank the families who participated in this study for sharing their samples and medical data, along with all the research groups who collaborate with the Broad CMG. G.L. was supported by the Fonds de recherche en santé du Québec (FRQS). A.S.J. was supported by a Massachusetts General Hospital (MGH) Fund for Medical Discovery Research Award. V.S.G. was supported by NIH NHGRI T32 (T32HG010464) and M.H.W. by NIH/NICHD K23HD102589. Sequencing and analysis were provided by the Broad CMG, funded by the National Human Genome Research Institute grants UM1HG008900, U01HG0011755 and R01HG009141. The content is solely the responsibility of the authors and does not necessarily represent the official views of the National Institutes of Health. Additional funding came from NIH/NINDS grants R01NS032457, R01NS058721, NIH/NIDDK grant RC2DK122397, Sanofi Genzyme, Ultragenyx, LGMD2I Research Fund, Samantha J. Brazzo Foundation, LGMD2D Foundation, Kurt+Peter Foundation, Muscular Dystrophy UK, Coalition to Cure Calpain 3, European Union’s Horizon 2020 research and innovation programme (grant agreement No. 779257 (Solve-RD)), the Murdoch Children’s Research Institute, the Harbig Foundation, the Victorian Government’s Operational Infrastructure Support Program, TUBITAK ((The Scientific and Technological Research Council of Turkey) Project No. 216S771), and Estonian Research Council grants PUT355, PRG471, PUTJD827, MOBTP175 and PSG774. We acknowledge the support provided by Samantha G. Beck, Yasmine Chahine, R. Sean Hill and Abbe Lai for ddPCR validation and variant interpretation. See supplemental for additional details.

## Web resources

seqr, https://seqr.broadinstitute.org/

GATK-gCNV, https://app.terra.bio/#workspaces/help-gatk/Germline-CNVs-GATK4

DECIPHER, https://www.deciphergenomics.org/

OMIM, https://www.omim.org/, https://genescout.omim.org/

ClinGen, https://search.clinicalgenome.org/kb/gene-dosage?page=1&size=25&search=

gnomAD, https://gnomad.broadinstitute.org/

ClinVar, https://www.ncbi.nlm.nih.gov/clinvar/

MatchMaker Exchange, https://www.matchmakerexchange.org

StrVCTVRE, https://strvctvre.berkeley.edu/

