## Supplemental for "Exome copy number variant detection, analysis and classification in a large cohort of families with undiagnosed rare genetic disease"

**Supplemental document**

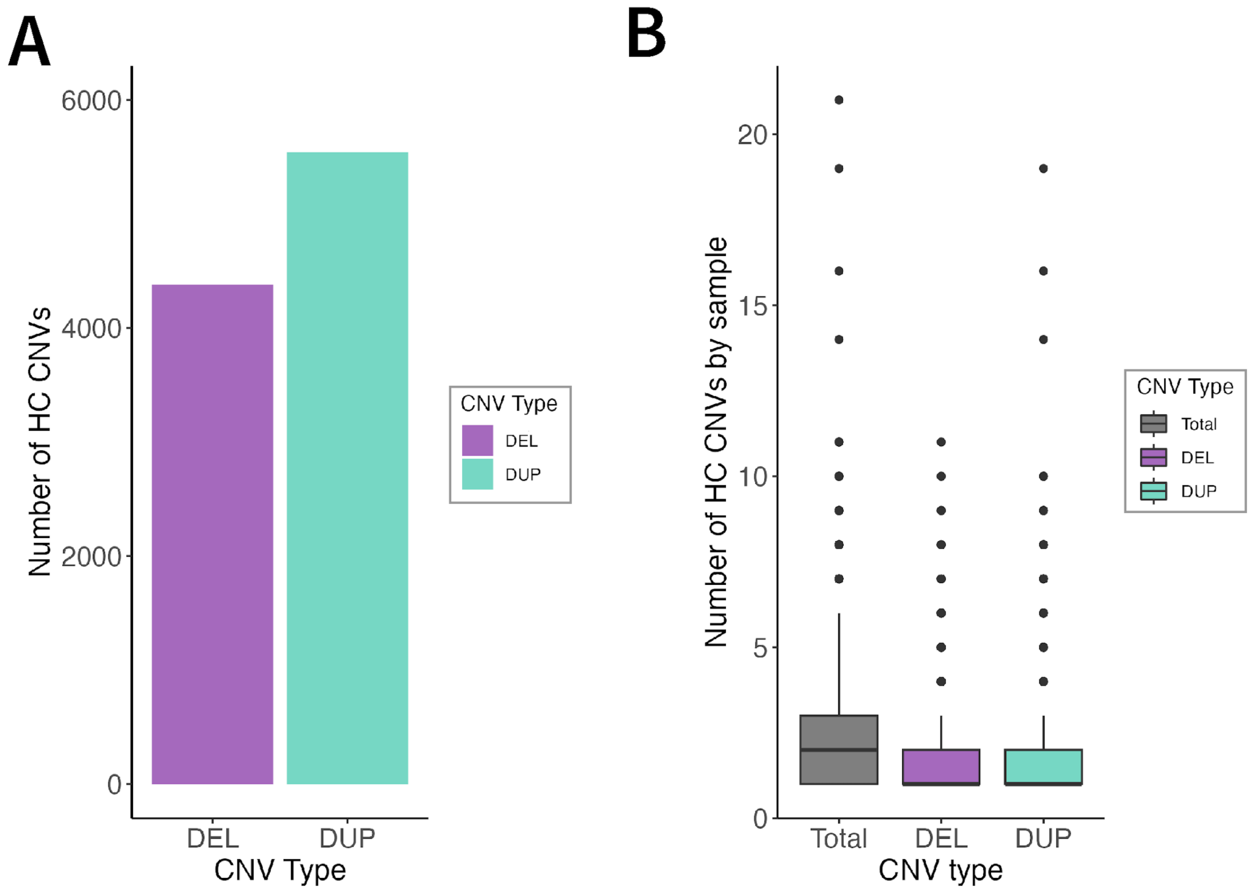


**Figure S1. Number of high-confidence CNVs across the Broad CMG cohort. A)** Number of high-confidence CNVs by type of CNV. Deletions are shown in purple and duplications in turquoise. A CNV is defined as high-confidence by GATK-gCNV (see Babadi *et al.*[1] for details) if 1) the CNV is present in a high-quality sample (with ≤ 200 autosomal raw CNV calls, of which at least 35 have QS >20); 2) the sample frequency of the call is ≤ 0.01 within the Broad callset; 3) the number of overlapped exons is ≥ 3; 4) the QS score is equal or greater than the QS threshold (QS>50 for duplications, >100 for deletions, and >400 for homozygous deletions). HC: high-confidence; QS: quality score; DEL: deletion; DUP: duplication. **B)** Boxplot showing the total number of rare (<1% frequency in the Broad CMG callset) high-confidence CNVs, deletions and duplications by individual in the Broad CMG cohort.

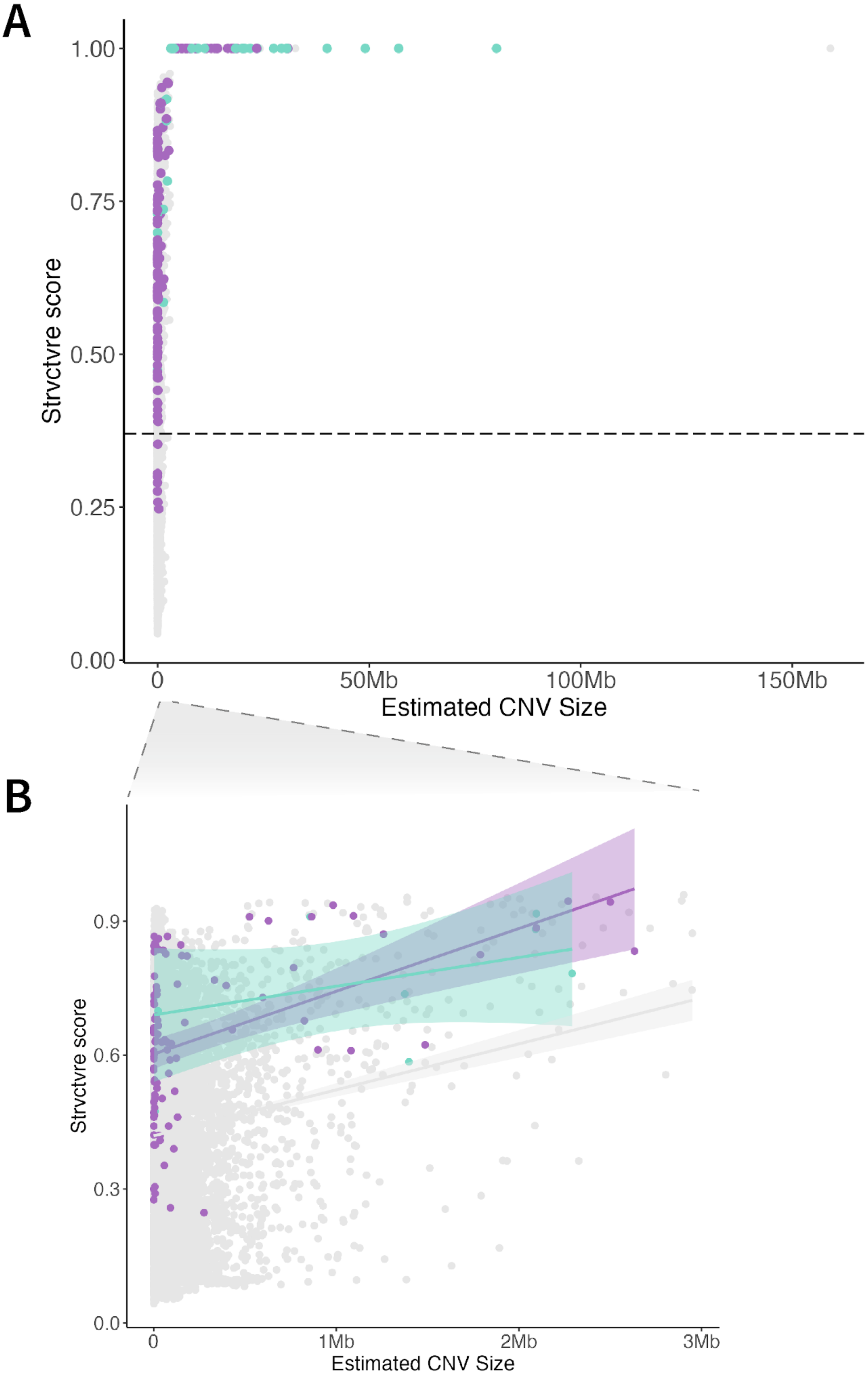
A
1.00
0.75
0.50
0.25
0.00
Strvctvre score
0
50Mb
100Mb
150Mb
Estimated CNV Size
B
0.9
0.6
0.3
0.0
Strvctvre score
0
1Mb
2Mb
3Mb
Estimated CNV Size

**Figure S2. Predicted StrVCTVRE score by estimated CNV size. A)** StrVCTVRE scores are shown by estimated CNV size. Only CNVs with annotated scores are represented. Non-causal variants are depicted in gray, causal deletions in purple and causal duplications in turquoise. Dashed horizontal line is set at StrVCTVRE score of 0.37, which value has been previously suggested as having a higher likelihood for a CNV of being deleterious.[2] We note that all CNVs with a size higher than 3 Mb have a StrVCTVRE score of 1. **B)** Zoomed panel of CNVs smaller than 3 Mb. Smoothed linear model lines are shown for non causal and causal deletions and duplications, with a positive correlation between StrVCTVRE score and estimated CNV size.

**Acknowledgements**
C.A.W. is an Investigator of the Howard Hughes Medical Institute and was supported by NIH/NINDS R01NS032457. C.G.B.'s laboratory is supported by intramural funds from NINDS. This research was supported by grants from the National Institutes of Health to F.H. (RC2DK122397) and E.H.S. (R01NS058721). MYO-SEQ was funded by Sanofi Genzyme, Ultragenyx, LGMD2I Research Fund, Samantha J. Brazzo Foundation, LGMD2D Foundation and Kurt+Peter Foundation, Muscular Dystrophy UK, and Coalition to Cure Calpain 3. A.T. has received funding from the European Union's Horizon 2020 research and innovation programme under grant agreement No. 779257 (Solve-RD). The Undiagnosed Disease Program Victoria (S.M.W, L.G., T.T., and Z.S.) acknowledges financial support from the Murdoch Children's Research Institute and the Harbig Foundation. The research conducted at the Murdoch Children's Research Institute was supported by the Victorian Government's Operational Infrastructure Support Program. Some patient samples were supported by TUBITAK (The Scientific and Technological Research Council of Turkey) Project No. 216S771. K.Õ. was supported by Estonian Research Council grants PUT355 and PRG471. S.P. is supported by Estonian Research Council grants PUTJD827, MOBTP175, PSG774.
